# State policies and clinicians’ and administrators’ perspectives on the inclusion of Medical Cannabis information in the Prescription Drug Monitoring Program

**DOI:** 10.1101/2025.07.24.25332003

**Authors:** Christina M. Gregor, Maria Y. Tian, Lorraine D. Tusing, Eric A. Wright, Brian J. Piper, Katrina M. Romagnoli

## Abstract

**Introduction:** Medical cannabis use is increasing worldwide, and information about its use could aid clinicians in decision making. A Prescription Drug Monitoring Program (PDMP) is an electronic database designed to track controlled substance (CII-CV) prescriptions; the use evolved to support patient care. We analyzed the perceived impact of cannabis information in the PDMP within an integrated health system in Pennsylvania and state PDMP policies around cannabis.

**Methods:** We conducted a sub-analysis of 50 semi-structured interview transcripts.

Interviews were conducted March-July 2023 with a multidisciplinary group of clinicians and administrators to understand cannabis use documentation. Twenty-four participants were asked if cannabis in the PDMP would impact their work. Additionally, we asked PDMP administrators of 50 US states and the District of Columbia (D.C.) if cannabis is in their PDMP. If yes, we inquired about software, timeframe, and data sharing.

**Results:** Almost two-thirds of 26 participants (N = 17, 65.4%) believed cannabis in the PDMP would positively impact patient care. Fifty states and D.C. replied to our survey. Six states (i.e., CT, LA, NY, OH, MS, and VA) have medical cannabis dispensations. Four states (i.e., AZ, IL, ND, and UT) have a medical cannabis card indicator in the PDMP.

Conclusions:

Participants perceive medical cannabis in the PDMP could enhance clinical decisions, but inclusion requires policy changes in 40 states and D.C. Rescheduling of cannabis could accelerate adoption of medical cannabis into the PDMP. The PDMP is an underutilized tool that could provide crucial information to clinicians, but substantial policy changes are necessary.

**Highlights:**

- Unrecognized drug interactions with cannabis can be harmful
- Minimal research exists on the impact on patient care with cannabis in the PDMP
- This research offers an examination of state legislation to patient care
- Clinicians suggest medical cannabis in the PDMP could benefit patient care

**Graphical Abstract.**
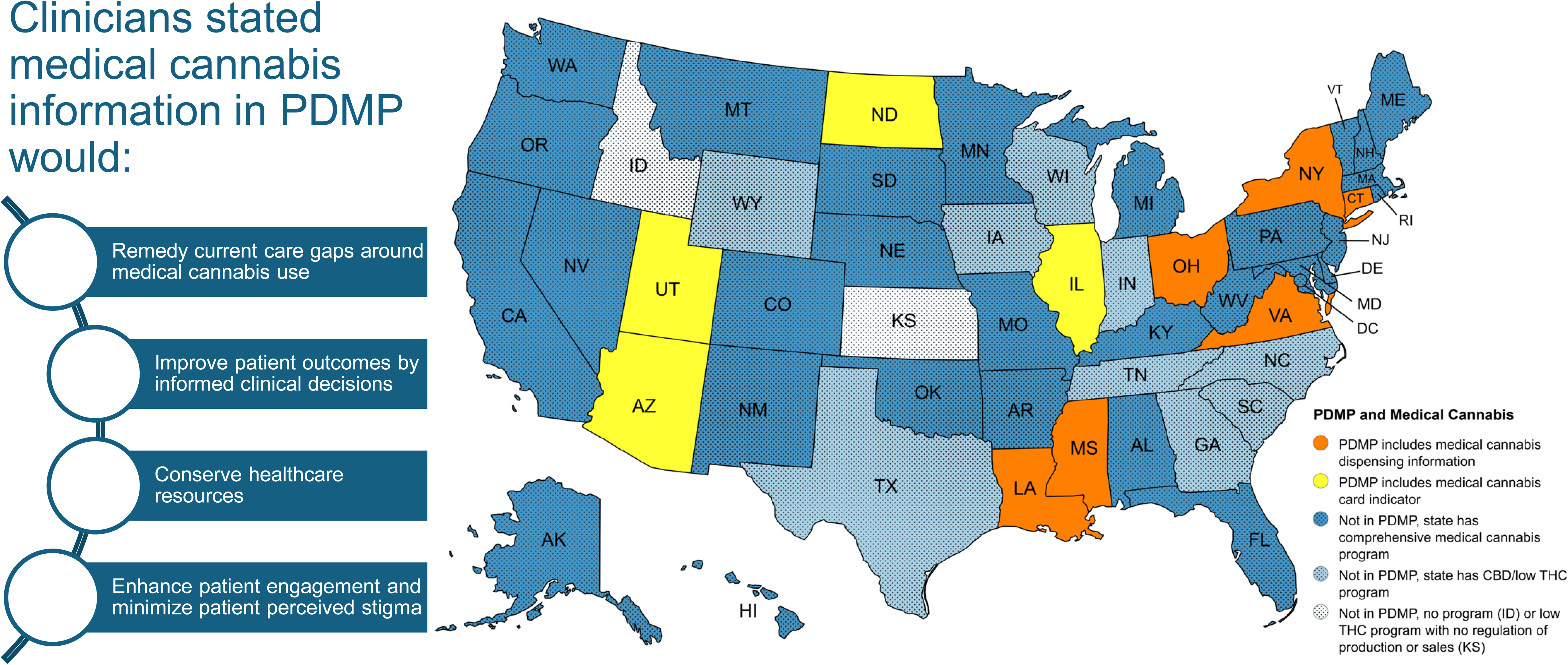
Themes from clinician interviews and heat map showing the ten states that have medical cannabis in their Prescription Drug Monitoring Program (PDMP) along with the characteristics of other states. Cannabidiol: CBD; Kansas: KS: Idaho: ID; Tetrahydrocannabinol: THC.

## Introduction

Patients are using cannabis at increasing rates to treat a wide array of health-related problems including chronic pain, HIV, cancer, Alzheimer’s Disease, and Parkinson’s Disease (Stains et al. 2025). The risk of developing cannabis use disorder is now 1 in 3, according to a 2022 national survey (Tang & Mccord, 2024). Another recent study found 11.7% of medical cannabis users developed cannabis use disorder (Cooke et al., 2023). Chronic cannabinoid users can develop Cannabinoid Hyperemesis Syndrome (CHS) which causes refractory nausea and vomiting that can lead to death (Nourbakhsh et al., 2019; Senderovich et al., 2022). At the time of this writing (7/2025), clinicians in the US do not prescribe medical cannabis, but rather follow the regulations of the state in which they are licensed for patients obtaining a medical cannabis card to acquire medical marijuana. In Pennsylvania, the role of a registered clinician (MD or DO) is to certify patients with at least one of the 24 qualifying conditions which would be benefited by treatment with medical marijuana (Commonwealth of Pennsylvania, 2024; Hirsch et al., 2024).

To the best of our knowledge, only minimal information about marijuana use is recorded in electronic health records (EHRs) (e.g., 4.8% medical use of cannabis is documented in the EHR compared 35.1% reported implicit medical use) (Lapham et al., 2022). At least one health system has a smart data element (SDE) implemented post medical cannabis legalization to track medical use of cannabis with relatively high completion rate by mostly rooming staff (i.e., certifying conditions (93.6%), product (92.9%), authorized dispensary (87.8%), active ingredient (83.3%), certifying provider (61.5%), and dosage (30.8%)) (Beiler et al., 2024). However, this is not comprehensive and does not solve the broader need for communicating this information across health systems. PDMPs offer a standardized way to track and monitor use (Center for Disease Control and Prevention., 2024) but only a few states have cannabis laws requiring cannabis to be part of the PDMP (Steuart, 2025).

PDMP use may support patient safety (Smith et al., 2023) by preventing drug abuse and diversion (Smith et al., 2023), adverse events (Senderovich et al., 2022) adverse drug interactions between cannabis/cannabinoids and warfarin, valproate, tacrolimus, and sirolimus (Nachnani et al., 2024)). PDMP use can also inform clinical decisions (Picco et al., 2021) and enable public health monitoring (Center for Disease Control and Prevention., 2024). Cannabis is not required to be tracked under U.S. federal law because it is a Schedule I substance, but states can decide to track this information. Presently, all PDMPs are regulated at the state level, and differ based on what substances are reported, how long data is retained, what software is used, and the ease of interface with health systems’ EHRs. Twenty-two states did not mention PDMPs in their medical cannabis laws (i.e., last update 2019) (Richard et al., 2021). However, eleven states require clinicians to review the PDMP before certifying patients for medical cannabis (Richard et al., 2021). An even smaller amount (i.e., two states) have laws that require dispensing data to be reported in the PDMP for medical cannabis (Richard et al., 2021). In Pennsylvania, clinicians are required to query the PDMP when initiating opioid therapy and every three months or more frequently when continuing opioid therapy to decrease diversion and abuse (Commonwealth of Pennsylvania, 2024; Dowell et al., 2022), but no such requirement exists for medical cannabis. At this time, Pennsylvania does not include medical marijuana use information in the PDMP.

In this study, we describe how the lack of state cannabis PDMP policies impact clinical care as perceived by clinicians within an integrated health system in Pennsylvania and identify areas of opportunity for improving the current state of medical cannabis use reporting. We also evaluated the current state of medical cannabis inclusion in the PDMPs of 50 US states and District of Columbia (D.C).

## Methods

### Study design

We conducted a cross-sectional mixed methods study using remote, qualitative semi-structured interviews of clinicians and administrators, and a survey of state employees responsible for their state’s PDMP.

### Study setting

All interview participants were employed in the same integrated health care system in PA at the time of the interviews. Surveys of PDMP administrators were sent to 50 states and the D.C.

### Study sample and data collection

#### Aim 1

We asked clinicians and administrators in an integrated health system in PA about their perceived value of having medical cannabis use information in the PDMP. This was part of a larger research study aiming to identify characteristics that influence EHR marijuana documentation. Initially, participants were not directly asked about their perspectives on cannabis use information in the PDMP, but it came up organically frequent enough that the study team decided to formally add the question to the interview guide. Therefore, some participants were not asked explicitly but discussed it anyway, while others were asked explicitly. Some early participants were not asked, and did not address it independently. We used purposive sampling to identify a Multidisciplinary Group of Clinicians (MGCs) (e.g., physicians, physician assistants, nurses, pharmacists, etc.), a Multidisciplinary Group of Clinician Administrators (MGCAs) (e.g., division chiefs, deans, directors, nurse supervisors, chiefs, coordinators, etc.) that practiced both as clinician and had an administrative role, and Multidisciplinary Group of Administrators (MGAs) (e.g., directors, assistant vice presidents (AVPs), operations managers, IT administrators, etc.) from various specialties and disciplines.

Participants were asked to discuss their experience with cannabis use information in the EHR, and if cannabis were added to the PDMP, how that might impact their work, if at all. All interviews were remote (i.e., Microsoft Teams, phone) and ranged between 30-60 minutes. Both trained staff members (AAC, LDT, AMP, BJP) and a qualitative research expert (KMR) used a semi-structured interview guide (see supplemental material) developed by the research team. Interviews occurred between March-July 2023. Interviews were recorded by calling into a transcription recording service on the phone, as well as a hand-held recorder as back-up. Participants did not receive any incentive for participation.

#### Aim 2

We sent the first questionnaire via email to understand which states had a medical cannabis requirement for the PDMP and the year it was enacted. State administrators who responded ‘yes’ received a follow-up questionnaire which inquired about the software used for their PDMP, what information is collected, the timeframe in which the information is collected (e.g., is it real time information or not), and if their information is shared with other states. We also asked if administrators could share a deidentified screenshot with test patient data of their PDMP user interface. We used the PDMP Training and Technical Assistance Center (PDMP-TTAC) to identify state PDMP administrator(s) (Institute for Intergovernmental Research, 2021). Additional emails were sent to administrators between September-October 2023 (Appendix A). These emails were re-sent to complete the data in February 2025 to five states whose administrators had not responded by October 2023. Phone calls were placed in February 2025 for nonresponses to three participants.

Of the state PDMP administrators who responded that their state includes cannabis in the PDMP, a second survey with additional questions was sent by email to inquire about their PDMP policies regarding cannabis (Appendix B). If they did not respond by email, a phone call was placed to collect responses.

### Ethics

Aim 1 and 2 were reviewed and approved by Geisinger’s Institutional Review Board (IRB # 2022-0498 (Human Subjects Research (i.e., interviews)) and IRB # 2023-1731 (Not Human Subjects Research (i.e., reach out to state administrators)).

### Data analysis

#### Aim 1

Semi-structured interviews were transcribed verbatim by an internal medical transcription team and deidentified by the study team. An initial deductive codebook was developed by LDT and was reviewed by the study team (KMR, BJP, CMG) prior to coding. We then used an inductive coding approach to add new codes. The transcripts were independently coded by two trained coders (CMG and LDT) and seven transcripts were double-coded to achieve inter-rater agreement of at least 75%. The rest of the coding was divided equally between the two coders. A quality check was completed on 25% of the remaining independently coded interviews. The study team reviewed 50 transcripts and coded the 32 transcripts that mentioned PDMP or similar program or were asked about PDMP explicitly. Twenty-four participants were asked the PDMP impact question (i.e., If you had access to marijuana usage information via the prescription drug monitoring program (PDMP) how would that impact your work?) to understand the impact of medical marijuana information in the PDMP. If we asked a variation of this question that did not call out impact or if they brought up the PDMP or similar program organically they were excluded from impact analysis but included overall in thematic analysis. We used reflexivity to determine this. Eighteen transcripts were excluded from analysis because there was no mention of PDMP or similar program in the content and they were not coded. This was because the inception of the idea about asking medical cannabis in the PDMP came after a participant mentioned that medical cannabis was analogous to narcotics. We then completed an IRB amendment to add the PDMP question. To capture demographics, we included an optional verbal survey at the end of the interview. Content and thematic analysis were used to analyze data.

#### Aim 2

States were categorized as: 1) those with medical cannabis dispensing information in the PDMP, 2) those where their PDMP has a medical card indicator but no dispense info, or 3) those without medical cannabis in the PDMP. A heatmap was prepared with MapChart.net (Figure 1). We analyzed perceptions of state policy impact on the health system through collecting responses from states with PDMP on their administrative and operative regulations surrounding their PDMP system. The responses were summarized in a table (Appendix C, Table A1).

**Figure 1.**
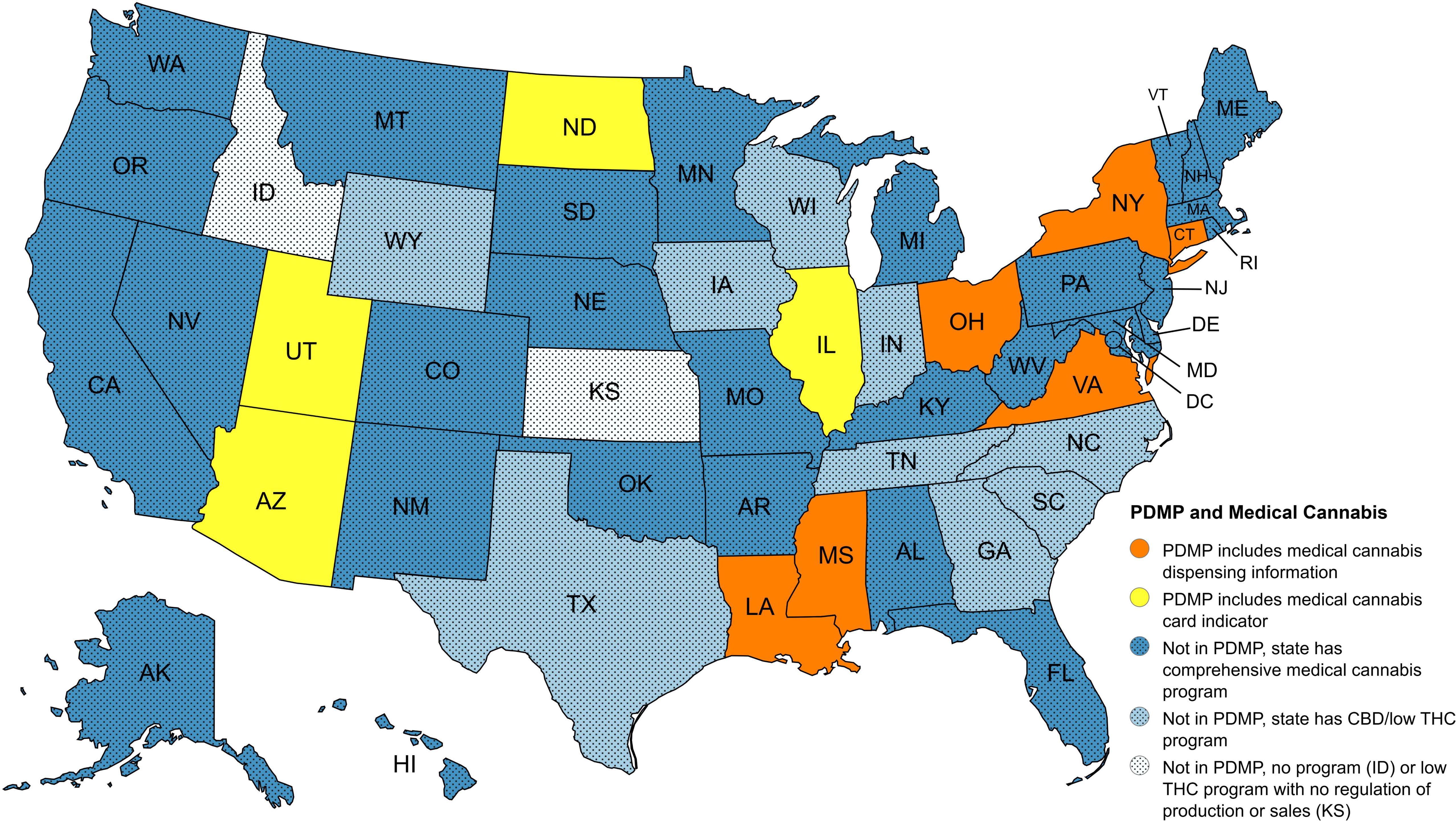
Heat map showing the ten states that have medical cannabis in their Prescription Drug Monitoring Program (PDMP) along with the characteristics of other states. Cannabidiol: CBD; Kansas: KS: Idaho: ID; Tetrahydrocannabinol: THC

#### Theoretical Framework

Team members continually discussed their perceptions of cannabis, the PDMP, and state policy in team meetings as well as the perception of the same topics among the participants. Once coding was finished and qualitative memos developed, the study team reviewed the results to address any biases (KMR, BJP, LDT, CMG).

## Results

### Demographics

Table 1 & 2 shows that of the 50 interviews, almost three-fifths of participants were female, a subset (12.0%) were non-White, 14.0% were primary care, one-third (32.0%) were physicians and one-quarter (26.0%) RN trained nurses.

**Table 1.**
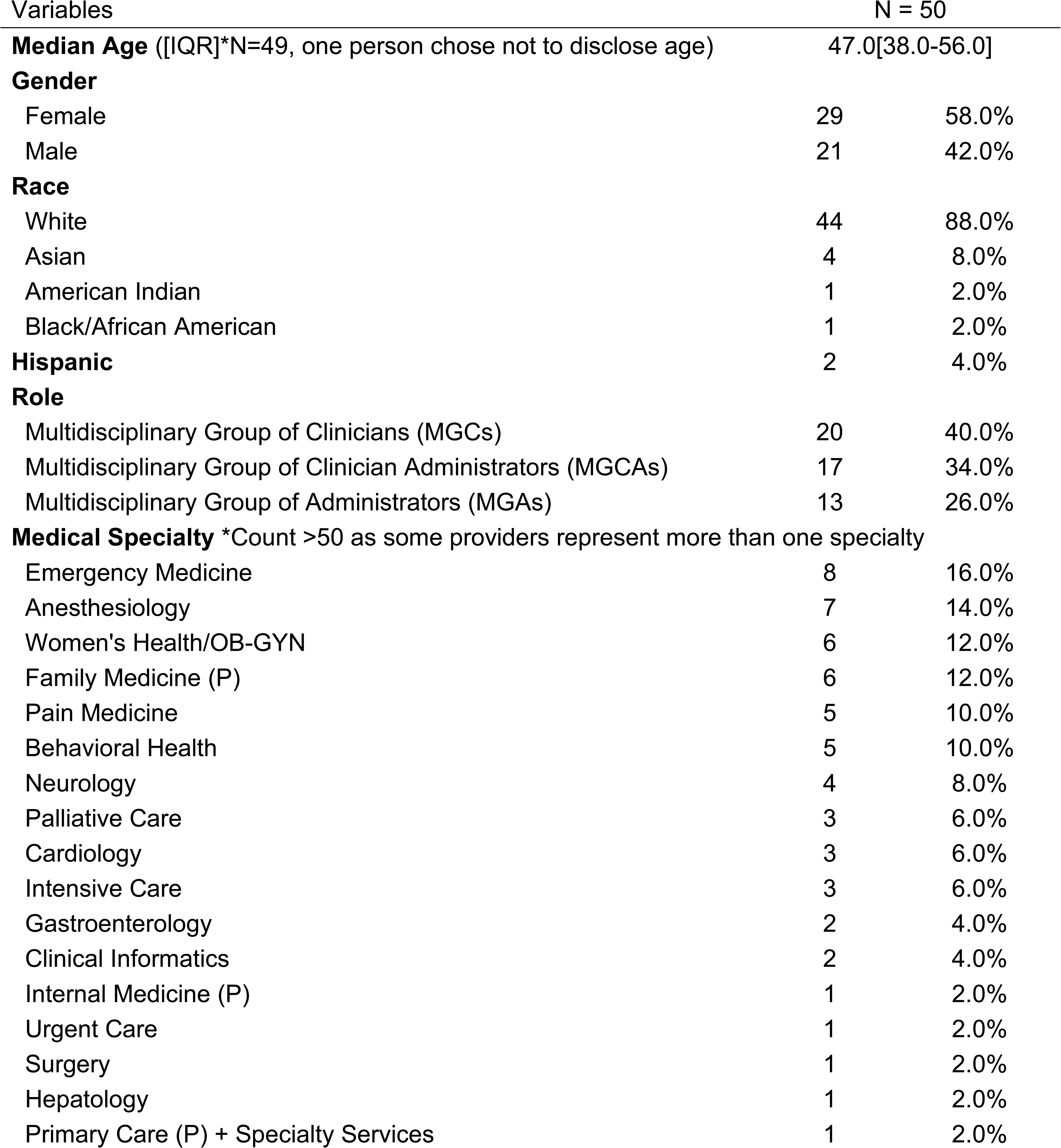
Demographics of 50 participants working in an integrated health system in Pennsylvania that completed a semi-structured interview about medical cannabis documentation.

**Table 2.** Demographics of 50 participants working in an integrated health system in Pennsylvania that completed a semi-structured interview about medical cannabis documentation.

| Variables | N = 50 |  |
| --- | --- | --- |
| **Credentials** * Count is >50 because some had more than 1 degree |  |  |
| RN | 13 | 26.0% |
| MD | 12 | 24.0% |
| Masters | 9 | 18.0% |
| DO | 4 | 8.0% |
| LPN | 4 | 8.0% |
| CRNP | 4 | 8.0% |
| BS/BA | 3 | 6.0% |
| PharmD | 2 | 4.0% |
| PA | 2 | 4.0% |
| CRNA | 2 | 4.0% |
| PhD | 1 | 2.0% |
| RPh | 1 | 2.0% |
| DNP | 1 | 2.0% |
| CNM | 1 | 2.0% |
| CMA | 1 | 2.0% |
| CPPM | 1 | 2.0% |
| **Time in years at Healthcare System** |  |  |
| < 5 years | 14 | 28.0% |
| 5-10 years | 14 | 28.0% |
| 11-20 years | 10 | 20.0% |
| >20 years | 8 | 16.0% |
| Not Asked | 4 | 8.0% |
| **Time in practice (N = 29)** |  |  |
| < 5 years | 3 | 6.0% |
| 5-10 years | 5 | 10.0% |
| 11-20 years | 10 | 20.0% |
| >20 years | 12 | 24.0% |
| Not Asked | 20 | 40.0% |

Twenty-six interview participants were not directly asked about cannabis use information in the PDMP. Of those 26 participants, eight participants addressed it organically. Two of the eight participants who addressed it organically also mentioned impact; they were included in the denominator for impact analysis (Figure 2). After the interview guide was updated and approved by the IRB, 24 additional interview participants were asked explicitly if having cannabis added to the PDMP would impact their work. Of the participants who were asked about the PDMP directly or brought it up organically and mentioned impact, almost two-thirds of them (n=17, 65.4%) believed the PDMP would positively impact their work.

**Figure 2.**
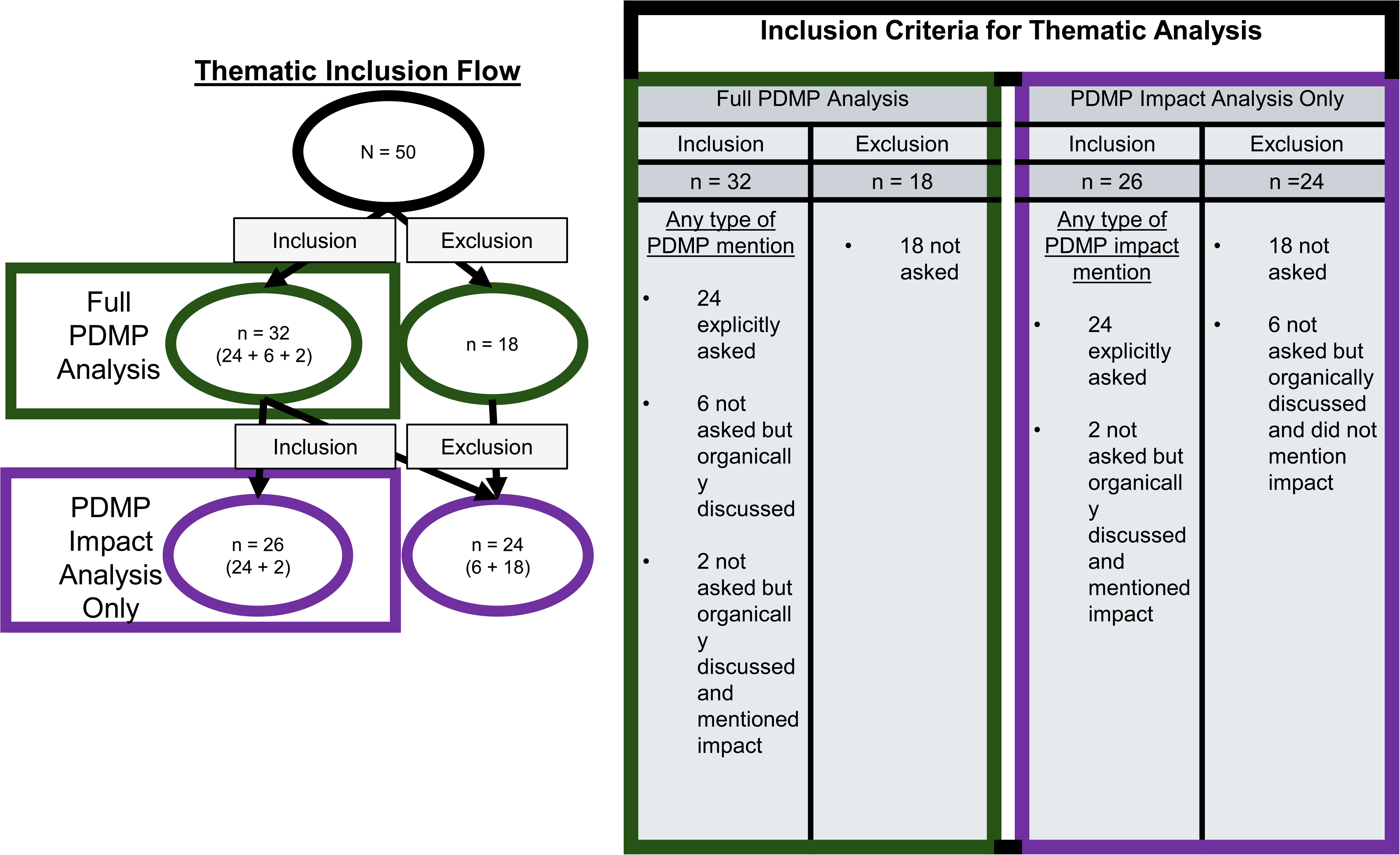
Depicts thirty-two individuals were included in the analysis for overall PDMP themes while twenty-six were included if they mentioned a type of impact organically or were explicitly asked

Twenty-five out of 26 were practicing clinicians. One administrator who did not practice clinically was asked the impact question in error. Eighteen participants were not asked any PDMP-related questions and did not bring up PDMP organically. They were excluded from the denominator of participants who discussed the PDMP either organically or were asked directly. Six people who brought the PDMP up organically but did not mention impact were also excluded from the denominator. Ten mentioned it would not impact their work because of one of the following: 1) does not affect clinical decisions in general or does not affect the treatment regimen for specific conditions they treat, 2) do not “prescribe” [sic] medical cannabis, or 3) did not state reason. Three mentioned it would not impact their work because PDMP review was not part of their role. Four mentioned it would negatively impact their work because it would add to their workload/frustration or could be punitive to clinicians.

### Interview Themes

*Medical cannabis policies play a key factor on how we deliver patient care, and changes in policy around medical cannabis in the PDMP offers opportunities to enhance patient care*.

The predominant theme was the current information gaps with respect to medical cannabis use information that might impact healthcare decision making. The PDMP may provide the opportunity to minimize the knowledge gap (Table 3). Other themes include having medical cannabis in PDMP may give time back to the clinician in investigating use in medical record (i.e., conserving healthcare resources), be an avenue to enhance patient engagement and minimize stigmatization, and an opportunity to improve patient outcomes with better informed clinical decisions. Participants indicate patients might feel less stigmatized if clinicians brought of the topic of cannabis based on what was found in the PDMP.

**Table 3.** Themes from about medical cannabis documentation with clinicians and administrators around patient care and Prescription Drug Monitoring Program (PDMP).

| Themes | Quotes |
| --- | --- |
| **<u>Remedy for current gaps in care</u>**<br>Gaps: rescreening, health literacy, self-report (i.e., reliability, social desirability bias), changes in use | “...a lot of patients don't bring it up...can't rescreen for everything at every follow-up appointment...they almost don't even think to bring it up because it's medical marijuana and maybe it's for pain or something else but not mental health-related, so they might not even mention it to me. So, I think if I had a PDMP to check, that would be awesome.” - *Psychiatrist & served as an Administrator* |
| **<u>Conserve healthcare resources</u>**<br>PDMP may conserve healthcare resources devoted to retrieval time of drug information from EHR | “... it's not based on the patient's word and it's just fairly documented...would be a good idea if it eventually gets federally decriminalized because the PDMP has saved a lot of practitioners a lot of researching and digging because it's very clearly laid out there...” - *Pain Medicine Physician* |
| **<u>Enhance patient engagement & minimize stigmatization</u>**<br>PDMP may enhance patient engagement and minimize stigmatization | “It would impact it significantly... when you start to go down the path now of asking about marijuana, it becomes many times, I can't say the majority of time, but a significant number of times a discussion of politics, right. I'm not asking somebody about why you smoke marijuana or how do you feel about this, it should be a national policy. It's a simple question of do you smoke marijuana, why you smoke marijuana, where's it coming from and how much...” - *Anesthesiologist & served as an Administrator* |
| **<u>Better patient outcomes</u>**<br>PDMP may provide better outcomes by informed clinical decisions around: drug/drug interactions, patient conversations, patient therapy plans, and suggestions for medical cannabis products | “Well, I think then it helps us make clinical decisions because...you already know most of the patient's relevant medical history before you get in there. ...having that information ahead of time can help facilitate that conversation around their substance use and also whether they're doing it legally or not.” – *Pharmacist*<br>“Why is this portion of supposed healthcare separated from all other healthcare from an information standpoint, so if the connectivity, the dispensaries have to gather all this data. They have to gather what they are giving them, how much, what they are using it for, what the dosage recommendations, what type of marijuana. All that stuff. Why in the world don't healthcare providers have direct access to all that information, electronically, so we know who they are. Because in my mind if we are saying this is a legitimate medical therapy, it ought to be treated like any other medication and we ought to have access to that information and then we could create electronic checks for drug interactions and we could understand the plan of therapy and what the goals were and all those pieces, because in today's world we are truly reliant on what the patient understands or remembers or is willing to share in order to get that information. So, but unfortunately it continues to ride the fine line between medical care and not and so we haven't applied the very standards we are trying to evoke on healthcare...”-*Administrator* |

Two people commented on the need for more education around cannabis to make recommendations such as what is typical dose and typical use, how to quantify a smokeless cannabis product vs smoked product before using the PDMP for patient care. One person mentioned being in a specialty where defining cannabis use beyond yes or no was not necessary without more education, unlike specialists in Anesthesia, Emergency Medicine, or Critical Care. One person commented that an algorithm was needed because he/she does not have time to log into PDMP and decipher what it means. This participant suggested an index should be developed to tell him/her exactly what to give a patient, how much and for how long. Another person commented that because clinicians do not “prescribe” [sic] medical cannabis, the PDMP may not be as helpful to influence medical cannabis use such as preventing patients from refilling it too early.

As observed in Figure 1, one fifth of all states (19.6%, 10 of 51 (states and D.C.)) and one fourth of states that have a comprehensive medical cannabis program (22.7.0%, 10 of 44 (states and D.C.)) have medical cannabis in their PDMP. PDMP programs with medical cannabis information included primarily exist in the eastern regions of the U.S. A few PDMP programs provide information about the patient having a medical cannabis card, but do not provide dispensing information in the west and Midwest regions. Five of the six states reported using Bamboo as their software to track medical cannabis. Some programs (Connecticut and Ohio) participate in interstate sharing of cannabis information but other states such as New York do not. The information availability time in all six states is 24 hours or fewer. While each of these states record medical cannabis transactions in their PDMP, the appearance and the type of information recorded varied from state to state (Appendix C, Table A1).

## Discussion

In this study, we described the impact on patient care with a state policy requiring medical cannabis to be in the PDMP. We also identified the states with a relevant medical cannabis policy, and the level of information shared with PDMP users. We learned clinicians at a rural health system in Pennsylvania perceive that medical cannabis drug information in the PDMP would provide useful information about medical cannabis use that may not exist in health systems nationwide without a medical cannabis PDMP policy. This information can provide an objective conversation starter for clinicians to use with patients and can be used to inform clinical decisions. After discussing with patients, clinicians can make recommendations about use.

An administrator summed up the current state of our health system around medical cannabis, “…it continues to ride the fine line between medical care and not and so we haven’t applied the very standards we are trying to evoke on healthcare”. As mentioned by an administrator, healthcare clinicians do not have access to the dispensaries’ medical cannabis drug information. This information could be helpful to create plan of therapy, goals, and electronic checks for drug interactions (Nachnani et al., 2024). By not having a national policy to report all medical cannabis drug information to health systems through the PDMP, states without a policy force health systems to develop their own solutions to capture and disseminate medical cannabis information such as an SDE. SDEs are an interim fix, but do not address the fundamental problem that this information depends on patient self-reporting to individual health care systems. Medical cannabis dispensaries, like pharmacies, produce and retain the most accurate information about what products are dispensed in what form, when, and to whom.

Effective medication management, patient safety, evidence-based medicine are three critical healthcare standards we aim to achieve, but a lack of national policies around medical cannabis prevents us from achieving these standards with respect to medical cannabis. A national policy does not exist requiring dispensaries to provide medical cannabis drug information. Details such as dosage, administration, indications, contraindications, warnings and precautions, adverse reactions, drug/drug interactions, and THC/CBD in a product are important to effective drug management. From our analysis, only four states captured the minimum (i.e., a discrete data points around cannabis use (i.e., yes/no)) while six states collected some clinically actionable information. Ten states out of the forty-four states with a comprehensive medical cannabis program collect cannabis information in the PDMP, but healthcare systems in those other thirty-four states may be collecting by other means such as SDEs in the EHR which rely on patients to self-report use and sharing that information via interoperable EHR systems. While a fraction of states participates in interstate sharing, a huge barrier still exists in both health management and access to health information if not all states are open to interstate sharing. The American Society of Addiction Medicine recommends all states report non-FDA-approved cannabis through the PDMP (American Society of Addiction Medicine, 2025). Also, a national policy may facilitate medical cannabis into the medical school curricula (i.e., in 2017 medical cannabis was a topic in only 9% of curricula in the United States) (Evanoff et al., 2017) which a few of interviewees mentioned they needed more education around cannabis to make recommendations.

The precise impact of medical cannabis on the value equation (i.e., Value = Patient Outcomes + Patient Experience / Direct Costs + Indirect Costs) is unclear. While fewer deaths and hospital readmissions occur with cannabis than opioids (Garnett & Miniño, 2024) a systematic review suggest legalized marijuana has shown reductions in opioid-related admissions (Okusanya et al., 2020). Clinicians’ perspectives from our interviews suggest having cannabis in the PDMP will improve patient outcomes with necessary information.

Currently, there is mixed evidence(Puac-Polanco et al., 2020; Tay et al., 2023) on the impact of PDMPs on population health. One systematic review suggests the reason for this mixed evidence is a lack of consistent robust methodology implementing the PDMP (Tay et al., 2023). Tay mentions the driving force around effectiveness of the PDMP was the uptake by end user (Tay et al., 2023). Some common cited barriers to PDMP review were usability, time constraints (Robinson et al., 2021; Smith et al., 2023; Tay et al., 2023), administrative burden, low perceived value of PDMP, and data integration (Robinson et al., 2021; Tay et al., 2023). Time constraints and administrative burden were also mentioned by participants in our study but only a few mentioned this compared to the majority mentioning positive impacts of the PDMP.

Potential reclassification of cannabis as Schedule III is unknown for 2025 (Veazey et al., 2025). However, if cannabis were to be rescheduled to Schedule III, medications are required to be entered into the PDMP (Owermohle, 2024), so this change would theoretically require medical marijuana use information to be reported via the PDMP, but it is not clear if that would happen. Marijuana was reclassified under the international Single Convention treaty in 2021 to Schedule I (*Legality of cannabis,* 2025). Some believe rescheduling to Schedule III does not violate the treaty because it will enhance the welfare of humankind and advancement of scientific research (Adlin, 2024). Rescheduling could offer the impetus to complete more clinical trials necessary for truly evidence-based medical marijuana treatment of qualifying conditions (FAAD, 2025). Discussion has occurred at a national level to remove cannabis from the schedule altogether. In that scenario, cannabis of any kind is unlikely to be included in any PDMPs (*Democrats urge Biden administration to deschedule marijuana,* 2024).

In our interviews, participants suggested a national policy about cannabis would help avoid perceived unnecessary political conversations between patient/clinician. The Code of Medical Ethics of the American Medical Association (AMA) recommends clinicians avoid political conversations, citing such concerns as imbalance of power between the patient and physician (American Medical Association, 2024). A clinician mentioned adding cannabis to PDMP in PA may resolve the need for clinicians to spend time playing “detective” for cannabis use in the medical record. Primary care physicians spend a median of 36.2 total minutes in the EHR before, during, and after each visit (Rotenstein et al., 2023). An SDE has been in place since 2017 for clinicians to document cannabis use manually in the EHR at the authors’ health system but it is used primarily by rooming staff (Beiler et al., 2024). This SDE requires manual user input of information, whereas the PDMP has been integrated within our EHR without user input, so clinicians do not have to enter other required information such as opioid prescriptions manually. Another study found the PDMP provided objective patient information (Picco et al., 2021). However, mixed evidence is available about clinician stigma towards patients with PDMP use. For example, practices sometimes remove from the practice “deceptive” patients who may be using prescription drugs for non-medical reasons (Picco et al., 2021). Also, patients with lower socioeconomic status tend to use emergency rooms at higher rates than other socioeconomic status groups, so their controlled substance histories would likely include more emergency clinicians, which could lead other emergency clinicians to stigmatize these patients and choose not to treat them (Picco et al., 2021). Decriminalization of cannabis may help with the underreporting and social acceptability/stigmatization of cannabis (National Academies of Science, Engineering, and Medicine, 2024). However, one report found the PDMP challenged prescribers to re-evaluate biases (Picco et al., 2021). It also found the PDMP helped clinicians conduct objective conversations with patients and helped detect patient dishonesty (Picco et al., 2021), which is similar to what participants in our study report about their use of the PDMP.

### Limitations

The findings in this Aim 1 may not be generalizable to other healthcare systems because we focused on one large, integrated healthcare system. Participants’ use of the PDMP might be different than other staff and clinicians at our health system. However, we recruited participants across many departments and roles to minimize this bias. We recognize cannabis is a politically charged topic. The cannabis PDMP question was asked in a way to decrease implicit bias by not asking how you feel about cannabis but how would adding cannabis use information to the PDMP impact your work. Although we used reflexivity as a main method of managing how researchers’ subjectively impacts the study, we also recognize the possibility of reflexivity bias (Walsh, 2003). Because we were limited to the questions we asked the states about cannabis integrated within their PDMP, there may have be further information in their policy we did not report as we did not examine each state’s policy directly. If and when cannabis policies are modified at state or federal level, additional research could be performed evaluating PDMP policies related to cannabis use information.

## Conclusions

The lack of access to patients’ medical cannabis use information is an information gap that could be addressed by using the pre-existing drug and medication use information sharing system (PDMP). State-managed PDMPs could ensure accurate and comprehensive documentation of medical cannabis use, akin to reporting of opioid prescriptions via the PDMP, thereby reducing reliance on patient self-reporting and incomplete manual data entry. Treating medical cannabis as a prescription drug and including that information in the PDMP could ensure clinically actionable dosage information is available. This could be achieved with a national policy for all the dispensaries to report medical cannabis transcriptions to the PDMP. Currently, the PDMP is an underutilized tool for medical cannabis in most states. The United States is able to create a national policy requiring medical cannabis to be treated as a prescription and requiring all states to include the information in the PDMP. This could optimize patient care in general and is specific to the potential for drug abuse.

## Supporting information

Supplemental Interview Guide

## Data Availability

The non-human subjects part of manuscript the data produced in the present study are available upon request to the authors, but the human subjects, qualitative interviews, part of this manuscript are not available upon request due to maintaining confidentiality to our participants.

## Glossary

CBD: Cannabidiol
CBD-A: Cannabidiolic Acid
CHS: Cannabinoid Hyperemesis Syndrome
CII-CV: Schedule II to Schedule V Controlled Substances
DOH: Department of Health
EHR: Electronic Health Record
IRB: Institutional Review Board
IQR: Interquartile Range
IT: Information Technology
MGAs: Multidisciplinary Group of Administrators
MGCAs: Multidisciplinary Group of Clinician Administrators
MGCs: Multidisciplinary Group of Clinicians
NDC: National Drug Code
PDMP: Prescription Drug Monitoring Program
SDE: Smart Data Element
THC: Tetrahydrocannabinol
THC-A: Tetrahydrocannabinolic Acid
THC/CBD: Tetrahydrocannabinol/Cannabidiol Ratio

## Acknowledgements

We would like to acknowledge the Geisinger Commonwealth School of Medicine (GCSOM) students: Alivia Roberts was involved in the data collection of the first round of PDMP survey inquiries and Courtney Chambers, Christina Shaffern, and Damien Balboa prepped the interview transcripts to be used in the analysis.

## CRediT authorship contribution statement

Christina M. Gregor: Conceptualization, Validation, Writing - Original Draft, Writing – Review & Editing, Visualization, Investigation, Formal Analysis, Data Curation. Maria Y. Tian: Conceptualization, Writing - Original Draft, Writing – Review & Editing, Investigation, Visualization. Lorraine Tusing: Conceptualization, Investigation, Supervision, Visualization, Writing - Review & Editing, Investigation, Funding acquisition, Project administration. Eric A. Wright: Conceptualization, Methodology, Validation, Supervision, Writing - Review & Editing. Brian J Piper: Conceptualization, Methodology, Validation, Visualization, Supervision, Investigation, Writing - Review & Editing. Katrina M. Romagnoli: Conceptualization, Methodology, Visualization, Investigation, Validation, Supervision, Writing - Review & Editing.

## Appendix A

Description: Questions we asked state administrators about the PDMP and marijuana

- Does your state PDMP collect information about marijuana?
- Is this information available to PDMP registrants?
- If yes, when (month/year) did your PDMP start doing this?

## Appendix B

Description: Follow-up questions we asked state administrators if they have marijuana in the PDMP

- What software (e.g., Bamboo, Canix, Distru, GrowFlo, MJ Freeway) do you use to track medical marijuana?
- Does it provide real-time information or is there a lag between sales and reporting? If not, what is the interval between sales and when it is reported in the PDMP? Is marijuana information shared with the PDMP’s of other states?
- What does the information look like in the PDMP? Is it by transaction? It is by dispensation? Does it provide any information about the route of administration? Is any summary information (THC/CBD) provided?

## Appendix C

**Table A1.**
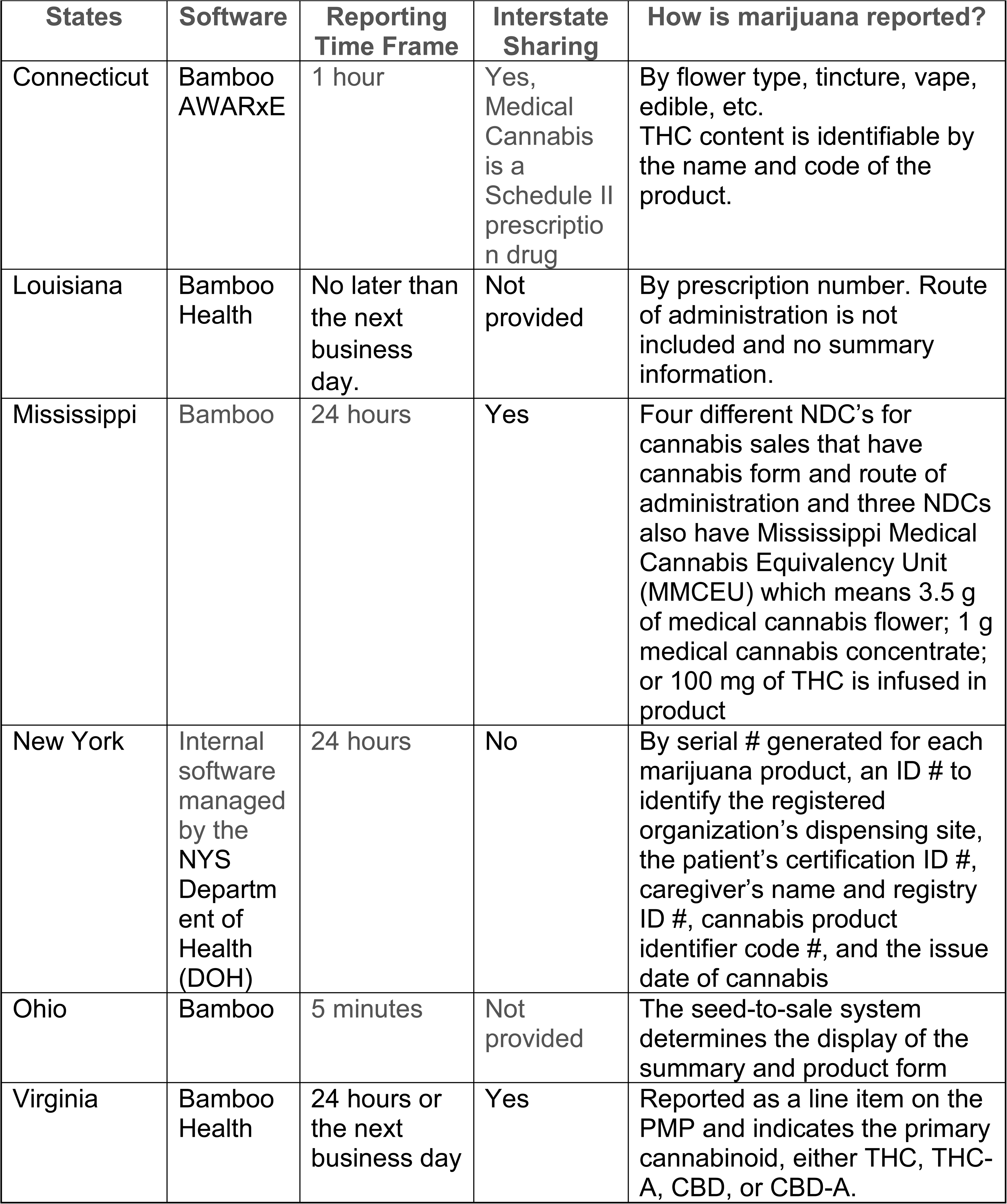
State Prescription Drug Monitoring Program (PDMP) open-ended responses about their medical marijuana policies including software, time frame, and data sharing. Cannabidiol: CBD; Cannabidiol Acid: CBD-A; National Drug Code: NDC; Tetrahydrocannabinol: THC.

