## Supplemental Interview Guide for "State policies and clinicians’ and administrators’ perspectives on the inclusion of Medical Cannabis information in the Prescription Drug Monitoring Program"

**Semi-structured Interview Guide**

**[Interviewer]:**

1. **First, I have some background questions for you.**
    - **Tell me a little about yourself**
    - Prompts: What is your role? How long have you been practicing? What kinds of conditions do you typically treat?
2. **Tell me about your experience with using information about marijuana usage as you treat your patients.**
    - *[if participant drifts from talking about information about marijuana usage to their opinions on marijuana policy, use of medical marijuana, etc.: thank them for sharing their thoughts, gently guide them back to the topic of interest. Example: thank you for sharing that perspective! In the interest of time, I want to focus on your experience with* ***using information about marijuana usage.*** *Can you tell me about your experience looking for and using information about marijuana usage in your patients?"]*
3. **Walk me through a typical patient for whom you look for information about marijuana usage in their EHR chart.**
   *[If they can't think of a "Typical" patient, ask about the most recent or most common patient]*
    - Prompt: What brings them into your clinic? What do you do first? Next? Next?
    - Prompt: what kind of information is valuable to you in collecting marijuana use information? Ex: do you want to know when they last smoked, how often, what they use it for, where they store it? What specific information is valuable for you as a clinician to hear about
    - Prompt: What are the resources that you use to help you through this process? (Examples: digital information resources, tools, people, services, etc.?)
    - Where in the record do you look for information about marijuana use?
    - How accessible/available is documentation of marijuana use in the EHR?
    - Prompt: Are there any other processes that you have not mentioned that are essential to the process of finding marijuana usage data in the EHR?
    - Prompt: What types of information is shared with the patient?
4. **Walk me through a typical patient who you end up recording in their EHR chart that they use marijuana.**
   *[If they can't think of a "Typical" patient, ask about the most recent or most common patient]*
    - Prompt: What brings them into your clinic? What do you do first? Next? Next?
    - Prompt: what kind of information is valuable to you in collecting marijuana use information? What specific information is valuable for you as a clinician to hear about? Ex: do you want to know when they last smoked, how often, what they use it for, where they store it, where they got it from?

        - If they can't think of a "Typical" patient, ask about the most recent or most common patient
    - Prompt: What are the resources that you use to help you through this process? (Examples: digital information resources, tools, people, services, etc.?)
    - Where in the record do you put information about marijuana use, if at all?
    - Prompt: is there ever a situation where you know a patient uses marijuana, but you decide not to record that information in the EHR? Why do you make that choice? How often do you experience this situation in a typical day?
    - Prompt: Are there any other processes that you have not mentioned that are essential to the process of inputting marijuana usage data in the EHR?
    - Prompt: What types of information is shared with the patient?

5. **Tell me about the most complex, difficult time you have ever experienced with a patient where information about marijuana usage was an important component to the care you were providing.**
   *[If they ask what you mean by difficult, turn the question back to them and ask them to decide what difficult means to them. Example: "I'm going to turn that question back to you. When you think of a difficult time, what do you think of?"]*
    - Prompt: What was difficult about it?
    - Prompt: What could have made it better?
6. **Tell me about the simplest, easiest you have ever experienced with a patient where information about marijuana usage was an important component to the care you were providing.**
    - Prompt: What made it easier than the most difficult experience you have had (people, tools, etc)?

7. **If you had access to marijuana usage information via the prescription drug monitoring program (PDMP) how would that impact your work?**

8. **If you had a magic wand that would make the process of inputting and using data about marijuana usage in the EHR better, what would you do? In other words, in the ideal world, where there were no limits on time or resources, what would information in the EHR about marijuana usage look like?**

9. **Is there anything else you want to tell me? Any questions you wish I had asked?**

### Demographic Questions

In these last few minutes, I will ask you some questions about yourself – please feel free to say “skip” if you would prefer not to answer.

**What was your sex at birth?**

☐ Female
☐ Male
☐ Intersex
☐ Other
☐ Choose not to disclose

**In terms of gender, how do you identify?**

☐ Female
☐ Male
☐ Female to Male transgender
☐ Genderqueer or non-conforming or non-binary or genderfluid
☐ Other
☐ Choose not to disclose

**What is your age?**

**Are you of Hispanic, Spanish-speaking, or Spanish origin?**

☐ Yes
☐ No
☐ Unknown/Prefer Not to Answer

**How would you describe your race (check all that apply)?**

☐ Black or African American
☐ White
☐ Asian
☐ American Indian or Alaskan Native
☐ Native Hawaiian or other Pacific Islander
☐ Other (Please specify): ____________________
☐ Prefer not to answer

**Thank you very much for your help. We know this is a difficult healthcare problem to address and appreciate your insights and advice very much. If you would like to see a summary of what we think we have learned from these interviews, we would be glad to provide it**
