## Supplemental Figure 1 for "State policies and clinicians’ and administrators’ perspectives on the inclusion of Medical Cannabis information in the Prescription Drug Monitoring Program"

Supplemental Figure 1. Provider views of cannabis record in Connecticut (A)

A. Connecticut's provider view of a patient marijuana record:

[redacted] | DOB: [redacted] | Sex: [redacted] | Report Prepared [redacted]



### Summary

| Summary | | Narcotics (excluding Buprenorphine) | | Sedatives | | Buprenorphine | |
|---|---|---|---|---|---|---|---|
| Total Prescriptions: | 971 | Current MME/day: | 0.00 | Current mg/day: | 0.00 | Current mg/day: | 0.00 |
| Total Prescribers: | 8 | 30 Day Avg MME/day: | 7.50 | 30 Day Avg mg/day: | 3.33 | 30 Day Avg mg/day: | 0.00 |
| Total Pharmacies: | 3 | Current Qty: | 0 | Current Qty: | 0 | Current Qty: | 0 |

### Prescriptions

| Fill Date | ID | Written | Drug | Qty | Days | Prescriber | Rx # | Pharmacy | Refill | Daily Dose* | Pymt Type | PMP |
|---|---|---|---|---|---|---|---|---|---|---|---|---|
| [redacted] | 3 | [redacted] | Durban Train Pure T441 21873 | 1.87 | 2 | [redacted] | [redacted] | [redacted] | 0 | | Private Pay | CT |
| [redacted] | 3 | [redacted] | Medical Marijauana Product | 1.71 | 2 | [redacted] | [redacted] | [redacted] | 0 | | Private Pay | CT |
| [redacted] | 3 | [redacted] | Yellow Haze Og Pure Vape T428 | 1.71 | 2 | [redacted] | [redacted] | [redacted] | 0 | | Private Pay | CT |
| [redacted] | 3 | [redacted] | Lemon Skunk Pure Vape T425 S 2 | 1.70 | 2 | [redacted] | [redacted] | [redacted] | 0 | | Private Pay | CT |
| [redacted] | 3 | [redacted] | Medical Marijauana Product | 1.71 | 2 | [redacted] | [redacted] | [redacted] | 0 | | Private Pay | CT |
| [redacted] | 3 | [redacted] | Double Bub T485 22522 | 1.94 | 2 | [redacted] | [redacted] | [redacted] | 0 | | Private Pay | CT |
| [redacted] | 3 | [redacted] | Select Elite Cartridge 1g Hawa | 3.51 | 2 | [redacted] | [redacted] | [redacted] | 0 | | Private Pay | CT |
| [redacted] | 3 | [redacted] | Lemon Skunk Pure Vape T425 S 2 | 1.70 | 2 | [redacted] | [redacted] | [redacted] | 0 | | Private Pay | CT |
| [redacted] | 1 | [redacted] | Hydrocodone-Acetamin 7.5-325 | 30.00 | 4 | [redacted] | [redacted] | [redacted] | 0 | 56.25 MME | Medicaid | CT |
| [redacted] | 3 | [redacted] | Motorbreath Pure Vape T444 H 2 | 1.77 | 2 | [redacted] | [redacted] | [redacted] | 0 | | Private Pay | CT |

This report shows marijuana products with assigned names and products with names not yet assigned. There is a seven-day delay period of when a product is approved and when it is updated in the PMP product list. This is captured in the program as the "Medical Marijuana Product" generic label of a recently purchased product. THC content is identifiable by name and code of product. T = % of mg or THC depending on product.
